# Molecular Mechanism of Parosmia

**DOI:** 10.1101/2021.02.05.21251085

**Authors:** Jane K. Parker, Christine E. Kelly, Simon B. Gane

## Abstract

The molecular stimuli that trigger a parosmic response have been identified. Parosmia is a debilitating disease in which familiar smells become distorted and unpleasant. Often a result of post infectious smell loss, incidences are increasing as the number of COVID-19 cases escalates worldwide. Little is understood of its pathophysiology, but the prevailing hypothesis for the underlying mechanism is a mis-wiring of olfactory sensory neurons. We identified 15 different molecular triggers in coffee using GC-Olfactometry as a relatively rapid screening tool for assessment of both quantitative olfactory loss and parosmia. This provides evidence for peripheral causation, but places constraints on the mis-wiring theory.

## Introduction

Prior to the COVID-19 pandemic, olfactory dysfunction was largely unrecognised, and often underestimated by health care professionals. Since the spread of severe acute respiratory syndrome coronavirus 2 (SARS-CoV-2), and the realisation that 50-65% of cases result in the loss of sense of smell (anosmia)^1^, there is a greater awareness of the debilitating effect of olfactory disorders^2^. Typically, in cases of COVID-19, normal olfactory function returns within a few weeks, but one study estimates 12% of all cases results in long term smell dysfunction^3^. With >100 million cases reported worldwide^4^ this is a significant problem facing the global population today.

Parosmia often occurs in the early stages of recovery from anosmia, typically 2-3 months after onset^1^, particularly in those whose anosmia was either acquired post-infection or post-traumatic brain injury^5^. Parosmia is a qualitative olfactory disorder in which familiar everyday smells become unpleasant and altered, to the extent that they become almost unrecognisable. These olfactory distortions are perceived in the presence of a stimulus. distinguishing it from phantosmia, where similar distortions are perceived in the absence of a stimulus. Those severely affected by parosmia find their quality of life deteriorates as everyday activities such as showering, oral care, eating and social interactions become a challenge. They report being distressed, scared and anxious about their future^6^ and, with many food aromas being intolerable, they start to reject food, leading to altered diets^7^, significant changes in weight, a decline in mental health and, in the most severe cases, to clinical depression^8-9^. The aim of this work was to gain insight into the mechanisms involved in parosmia. In 2013, coffee and chocolate were found to elicit distorted olfactory experiences in parosmia^10^ and more recently, coffee, meat, onion, garlic, egg, mint/toothpaste were identified in a thematic analysis of group posts on social media^6^. These foods contain aroma compounds with some of the lowest odour-thresholds known, and we suggest that these compounds may be involved in triggering olfactory distortions. Our original hypothesis was based on the mechanisms underlying parosmia being at least partly related to widespread loss of olfactory neurons (OSN). We proposed that as OSNs regenerate from basal stem cells, selective detection of just the pungent highly odour-active compounds might result in a distorted perception of certain foods and beverages. Whether this would be sufficient to cause the strong sense of disgust often reported with parosmia, was not clear. For those with a normal sense of smell, the perception of these potent molecules is moderated by tens of other less odour-active aroma compounds, acting synergistically to produce the desirable balanced characteristic aroma of coffee as we know it.

Our approach is novel in that we use GC-Olfactometry (GC-O) to determine which of the aroma compounds present in the headspace of coffee are responsible for the distortions and the sense of disgust experienced by parosmic participants. Gas chromatography separates the hundreds of volatile components present in the headspace which, when coupled to an odour-port, allows subjects to sniff and assess each component as it elutes from the column, thereby allowing us to screen multiple aroma compounds in a short time.

## Results and Observations

### Participants

Table 1 shows demographic data for all participants (N = 45) with full details provided in Supplementary Table S1. All participants were non-smokers and self-reported that they could taste the difference between salt and sugar. Aetiology of parosmia was post infection for 29/30 those with parosmia, with one case of traumatic brain injury. Pre-COVID-19 parosmic and non-parosmic individuals were age matched with mean ages of 56 and 49 y respectively and no significant difference between the two groups (p=0.12), whereas post-COVID-19 participants were significantly younger (mean age 37 y) than their pre-COVID-19 counterparts or the non-parosmic group (p<0.000, p=0.008 respectively).

**Table 1.** Summary of Participant Demographics

| Participant demographic data | No | Male | Female | White | Mean<br>age | Age<br>range | CRS | PI | TBI |
| --- | --- | --- | --- | --- | --- | --- | --- | --- | --- |
| All Participants | 45 | 12 | 33 | 43 | 53 | 23-73 | 3 | 29 | 1 |
| Pre-COVID-19 parosmics | 15 | 3 | 12 | 15 | 56 | 33-73 | 1 | 14 | 1 |
| Post-COVID-19 parosmics | 15 | 3 | 12 | 14 | 37 | 19-60 | 1 | 15 | 0 |
| Non-parosmics | 15 | 6 | 9 | 14 | 49 | 33-71 | 2 | na | na |

CRS = chronic rhinosinusitis, PI = post infection, TBI = post traumatic brain injury, na = not
applicable

### Olfactory Function

Bilateral olfactory function was assessed using the complete validated orthonasal psychophysical Sniffin’ Sticks test (Burghart, Wedel, Germany)^11^ based on the threshold of phenylethyl alcohol (T), discrimination (D) and identification (I) tests which gives a TDI score (0-48). TDI scores were significantly lower in pre- and post-COVID-19 participants (mean 27 and 28 respectively) compared to the non-parosmics group (mean 37) (ANOVA, p<0.0001 for both groups) but there was no significant difference between pre- and post-COVID-19 parosmic individuals (p=0.71). The TDI scores of the combined parosmic groups ranged from anosmic to normosmic (10-38). Ten of this group were classified as normosmic on raw TDI score, increasing to 17 (more than half of the group) when age adjustment was applied^12^. On the contrary, three parosmic participants scoring <16 were classified as functionally anosmic. We demonstrate here that although on average most parosmic participants had a low olfactory function, parosmia also occurs in those with a normal olfactory function.

### Gas Chromatography-Olfactometry

The number of aromas detected by GC-O by non-parosmic participants was significantly higher than for pre- or post-COVID-19 parosmic participants (p<0.0001) (means 37, 19 and 19 respectively). This parameter correlated well with the TDI score (R^2^=0.65, Fig. 1A). However, the mean number of molecular triggers detected by parosmic participants was only 6.4 (range 0-13) indicating that on average parosmic participants detected twice as many “normal” aroma molecules compared to trigger molecules. This has never been demonstrated before and shows that, in those presenting with parosmia, some OSNs are functioning normally and it is only specific OSNs (or combinations of them) which trigger the altered perception of food and the sense of disgust. There was no strong correlation between the number of molecular triggers reported and TDI score (R^2^=0.16, Fig. 1B) suggesting that although quantitative and qualitative olfactory disorders may occur together, their mechanism may be quite different.

**Fig. 1.**
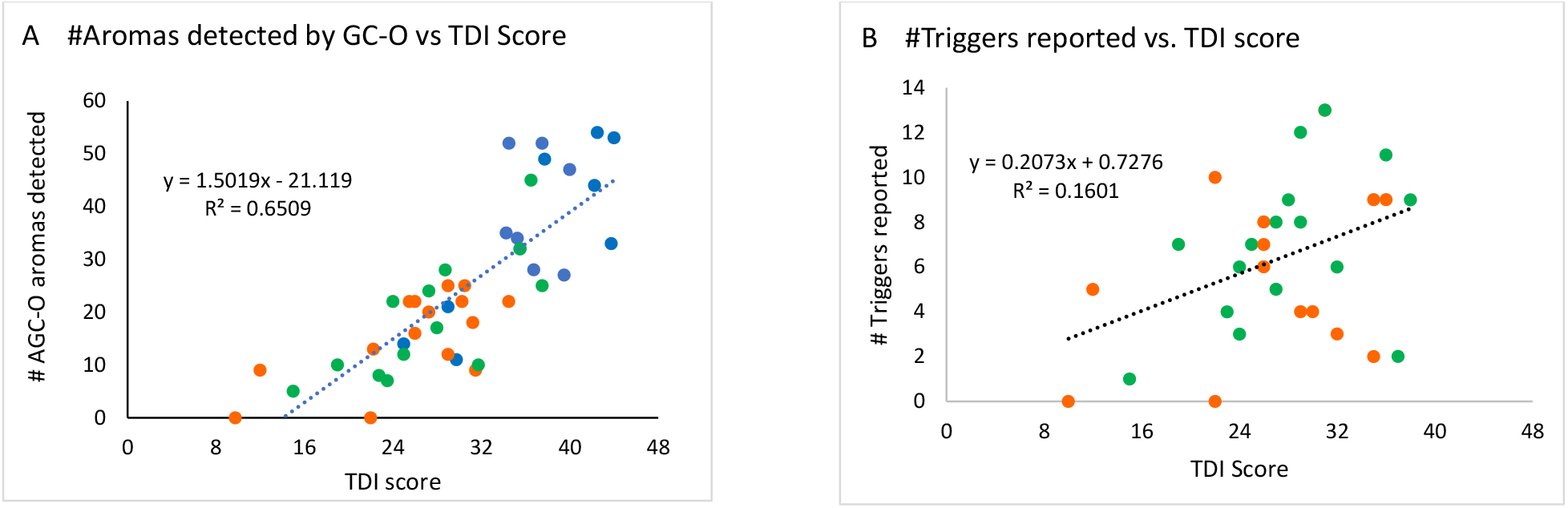
Correlations between olfactory function and GC-O. A: Correlation between TDI score and number of GC-O aromas detected at the GC-O. B: Relationship between number of triggers detected in the coffee extract and TDI score. In both figures non-parosmic participants = blue, pre-COVID-19 parosmic participants = orange, post-COVID-19 parosmic participants = green.

### Molecular triggers

Molecular triggers identified in coffee headspace by >2 parosmic participants are shown in Table 2. The most frequently reported trigger (21/30) is 2-furanmethanethiol. It imparts a roasted coffee-like odour and has a particularly low odour threshold in water of 0.004 ug/kg^13^. Whereas the non-parosmic participants used a range of food related terms to describe it (including “coffee”, “roasty”, “popcorn”, “smoky”), it was commonly described by parosmic participants as “new coffee” (relating to the altered smell of coffee since onset of parosmia), or with words describing its hedonic quality (“disgusting”, “repulsive” and “dirty”). All non-parosmic participants except one non-coffee drinker detected this compound. Five parosmic participants with low TDI scores did not detect this compound but, when assessing a 100-fold more concentrated coffee extract by GC-O, described it as a trigger. Four parosmic participants described it in the same way as non-parosmic participants (“biscuit”, “toasty” or “roasty”) indicating that it is not universally parosmic, but certainly an important and frequent molecular trigger of parosmia. The isomeric, and equally potent 2-methyl-3-furanthiol (threshold 0.0004 ug/kg in water^14^) and its corresponding methyl disulfide were also detected, but reported less frequently as triggers. They are character impact compounds in meat, and we confirmed in four parosmic participants who assessed grilled chicken by GC-O that these compounds also triggered parosmia in meat.

**Table 2.** Molecular triggers reported in coffee by at least 3 pre- and post-COVID-19 parosmic participants

| Molecular triggers | Code | Class | Odour threshold<br>ug/L | Number times<br>detected<br>by parosmic | Number times<br>reported<br>as trigger | Source<sup>a</sup> |
| --- | --- | --- | --- | --- | --- | --- |
| 2-furanmethanethiol | fft | furanthiol | 0.005<sup>13</sup> | 24 | 20 | TCI |
| 2-ethyl-3,6-dimethylpyrazine | edmp1 | pyrazine | 0.01 | 19 | 14 | Oxford |
| 2,3-diethyl-5-methylpyrazine | demp | pyrazine | 0.05<sup>13</sup> | 21 | 12 | Oxford |
| 2-furanmethyl methyl disulfide | ffssm | disulfide | 0.04<sup>16</sup> | 18 | 11 | Sigma |
| 2-methyl-3-furanthiol | mft | furanthiol | 0.0004<sup>14</sup> | 20 | 10 | TCI |
| 2-ethyl-3,5-dimethylpyrazine | edmp2 | pyrazine | 1<sup>37</sup> | 19 | 10 | Oxford |
| 2-methyl-3-furyl methyl disulfide | mfssm | disulfide | 0.004<sup>14</sup> | 19 | 10 | Sigma |
| 3-methyl-2-butene-1-thiol | mbt | thiol | 0.005<sup>Aroxa</sup> | 21 | 9 | Aroxa |
| 2-ethyl-3-methoxypyrazine | emp | methoxypyrazine | 0.4<sup>16</sup> | 12 | 9 | Sigma |
| 2-isobutyl-3-methoxypyrazine | ibmp | methoxypyrazine | 0.002<sup>13</sup> | 18 | 7 | Sigma |
| 3-mercapto-3-methylbutanol | 3mmbo | thiol |  | 14 | 6 | Sigma |
| sotolone | sot | furanone | 0.5<sup>13</sup> | 10 | 6 | Sigma |
| trimethylpyrazine | tmp | pyrazine | 9.6<sup>16</sup> | 11 | 5 | Sigma |
| guaiacol | gc | phenol | 12<sup>13</sup> | 15 | 5 | Sigma |
| 3-mercapto-3-methylbutyl formate | 3mmbf | thiol ester |  | 15 | 5 | Fluorochem |
| unknown LRI 981 | unk |  |  | 12 | 4 | - |
| 2-isopropyl-3-methoxypyrazine | ipmp | methoxypyrazine | 0.001<sup>13</sup> | 15 | 3 | TCI |

<sup>a</sup>Source of authentic standard: Aroxa (Leatherhead, UK), Fluorochem (Hadfield, UK), Oxford Chemicals (Hartlepool, UK), Sigma (Poole, UK), TCI (Oxford, UK).

2-Ethyl-3,6-dimethylpyrazine was the second most frequent trigger in coffee (14/30 times), described with a variety of food terms by non-parosmic participants, but by “new coffee”, “unpleasant” and “distorted” by parosmic participants. Other trisubstituted pyrazines (2,3-diethyl-5-methylpyrazine, 2-ethyl-3,5-dimethylpyrazine and trimethylpyrazine) were common triggers. These pyrazines are highly odour-active compounds in roasted, fried and baked goods, and we confirmed by GC-O that these compounds also triggered parosmia in cocoa (N=4), grilled chicken (N=4) and peanut butter [N=3]. 2-Ethyl-3-methoxypyrazine, 2-isobutyl-3-methoxypyrazine and 2-isopropyl-3-methoxyprazine were common triggers in coffee, and we confirmed that these also contributed to the parosmic character of bell peppers (N=5), where they are character impact compounds.

The furanthiols, disulfides and the pyrazines which represent 9 of the 10 most frequently reported trigger molecules, are interesting as they are all potent heterocyclic molecules containing an exposed heteroatom with relatively good Lewis base properties.

Another important trigger, 3-methyl-2-butene-1-thiol, with a pungent weedy character and low threshold (0.0002 ug/L), was reported as a trigger 9/30 times. Although not heterocyclic, it contains the same α,β-unsaturated thiol moiety as 2-furanmethanethiol. The thiol ester, 3-mercapto-3-methylbutyl formate, is one of the most potent aroma compounds in coffee^15^ and was the most frequently detected by parosmic participants (15/30), but only reported 6 times as a trigger. Although thiols and disulfides seem to be effective triggers for parosmia, there are two notable exceptions. Methanethiol (odour threshold 0.02 ug/L^16^), which was detected by some non-parosmic participants, was not detected by any of the parosmic participants, even in 100-fold concentrated coffee. Likewise, dimethyl trisulfide (0.01 ug/L^13^) is an exceptionally potent compound detected by 12/15 non-parsomic participants but only by 4 parosmic participants, and only once reported as a trigger.

Furthermore, a handful of compounds were occasionally detected by but never reported as triggers. (*E*)-β-Damascenone, a key odour-active compounds in coffee with a low odour threshold (0.01 ug/kg^13^), was detected by 6 parosmic participants and always described as jammy and fruity. Likewise, 4-ethylguaiacol was detected by 7 parosmic participants and always described as spicy, sweet and smoky, but never parosmic.

### Cluster analysis

Agglomerative hierarchical cluster analysis was carried out on the combined intensity data from all parosmic participants of all potential triggers reported >3 times. It showed four significant clusters of compounds (Fig. 2). A structure activity pattern starts to emerge, suggesting, for example, that some participants might perceive thiols more intensely and others may perceive pyrazines more intensely. When the same analysis was carried out using the data from the non-parosmics, the same clusters did not emerge, but N was small, and we cannot draw any conclusions from this. It is likely that in the general population, there are significant differences in the relative perception of these compounds.

**Fig. 2.**
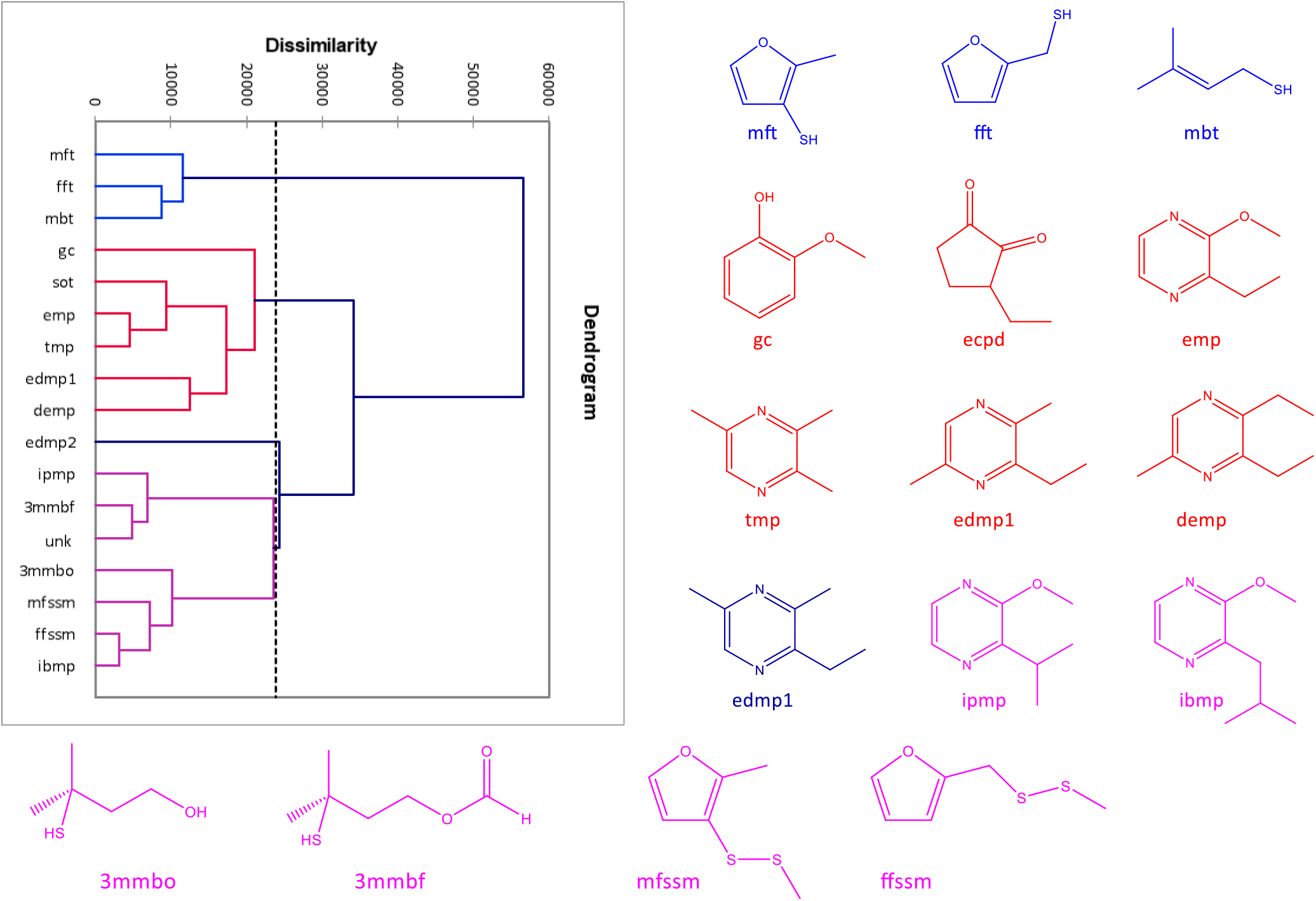
Agglomerative Hierarchical Clustering of intensity scores for 17 most frequently reported triggers

### Correlation between ligand structure and odour receptor (OR)?

Identifying a small number of common molecular triggers for parosmia raised the obvious question of an olfactory receptor similarity. To determine whether the clusters are associated with any of the known ligand odour receptor pairs, we searched the ODORactor database^17^. We found no obvious segregation of triggers by olfactory receptor (Fig. 3). Most of the triggers activated (with > 50% probability) either OR1G1 or OR52D1. We then compared molecules never reported as triggers such as disubstituted pyrazines, indole, skatole, cresol and found these to activate the same ORs, making it unlikely that these olfactory receptors are the source of the parosmic signal. OR1G1 is known to be very broadly tuned and bind odorants of different chemical classes^18^. However, only a fraction of the known olfactory receptors have been deorphaned, and further identification of ligand-OR pairs is required.

**Fig. 3.**
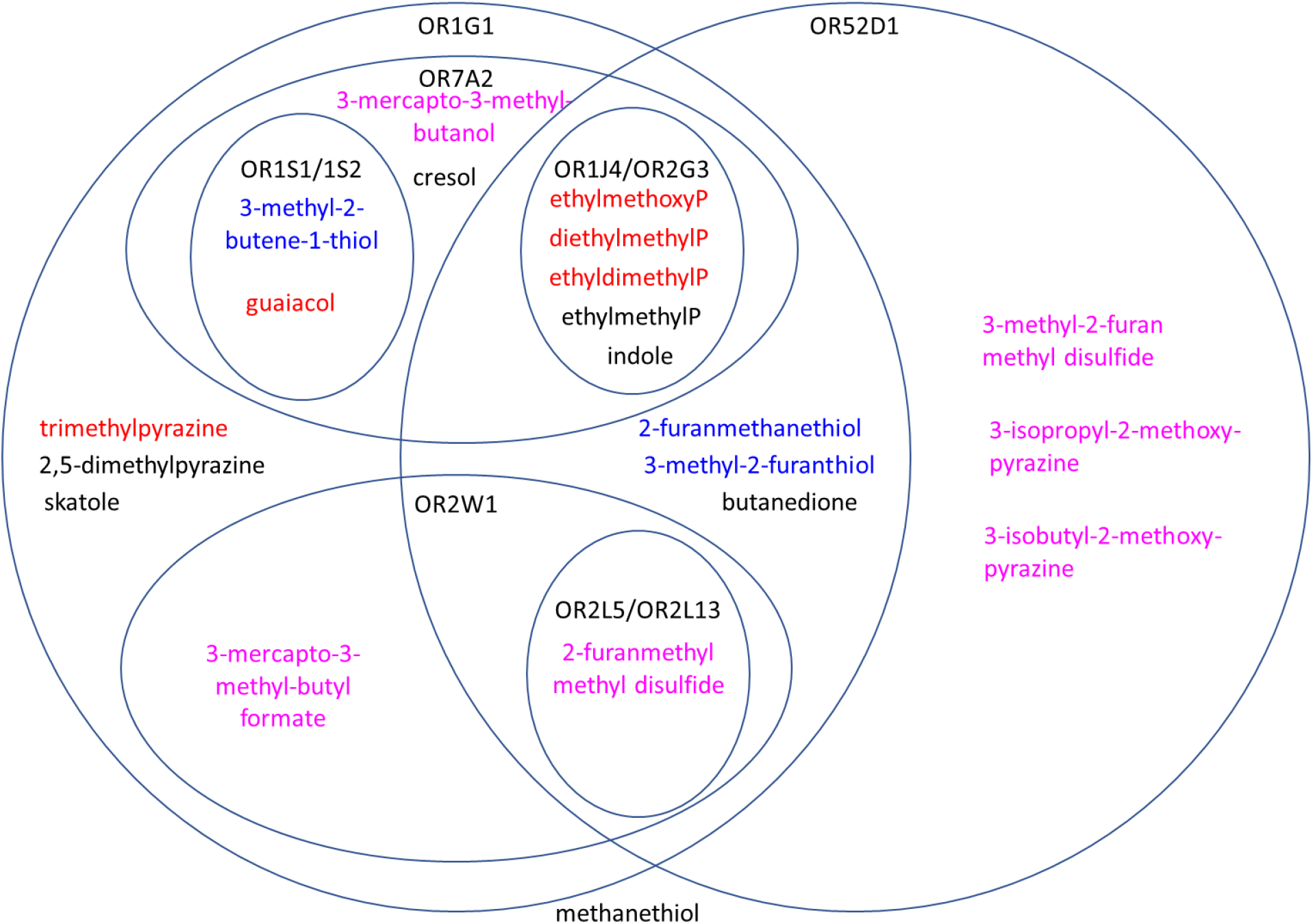
Venn diagram showing molecular triggers and their known odour receptors. ORs are blue, compounds in red are known triggers, whereas compounds in black have not been identified as triggers. 2-Ethyl-5-methylpyrazine and 2,5-dimethypyrazine may be triggers at much higher concentrations but are rarely odour-active (high odour thresholds), other research (unreported) suggests indole and skatole are unlikely to be triggers. For OR1J4 anOR2G3, pyrazines have been abbreviated with a P: triggers within OR1J4 and OR2G3 from top to bottom are 3-ethyl-2-methoxypyrazine, 2,3-diethyl-3-methylpyrazine, 2-ethyl-3,5-dimethylpyrazine, and non triggers 2-ethyl-5-methylpyrazine and indole. In addition, the following ligand OR pairs were retrieved: butanedione: OR6Y1; guaiacol: OR5L2, OR1L8, OR5AS1, OR8G2, OR4K15, OR5D18, OR10R2, OR5V1, OR8J1, OR6C75, OR1F1, OR8H2, OR1J2, OR7G1, OR1E3; 3-methyl-2-butene-1-thiol: OR1L3; methanethiol: OR5B12, OR5AR1, OR4S2, OR2W5, OR5M3.

### Faecal odours

Parosmic participants often comment that the smell of faeces is never as unpleasant as before, often smelling like other parosmic food, or even more pleasant and biscuity^6^, presenting the interesting corollary that foods smell of faeces yet faeces smell of food. One normosmic and two parosmic researchers carried out GC-O on a 50% faecal slurry in water. Whilst the normosmic scored the intensity of the volatiles normally associated with faeces (indole, skatole and p-cresol^19^) as close to strongest imaginable, the parosmic researchers were unaware of these foul smells, rather reporting many of the potent triggers already mentioned in coffee. This provides a neat explanation as to why the changes in valence for faecal samples is reversed. In the absence of signals from the usual faecal compounds, parosmic participants detect other potent volatiles in the sample, normally masked by the faecal compounds for normosmics, which may elicit any number of odour percepts depending on the sensitivity of the parosmic to the other compounds present.

### Participant observations

i. A case of parosmia not preceded by anosmia: One participant identified 45 aromas, of which just 2 were triggers. The more intense one was 2-ethyl-3,5-dimethylpyrazine and the second a mild and unidentified compound. They had a TDI score of 37, had tested positive for COVID-19 antibodies but reported no loss of sense of smell. This is an unusual case where parosmia was not preceded by anosmia.
ii. A case of parosmia improving with no concomitant increase in olfactory function: After 4 months one participant showed no improvement in threshold score, a decrease in the number of GC-O aromas detected (22 to 16), and a 5-fold decrease in intensity scores (750 to 147) yet reported an improvement in parosmia. Further work on the temporal aspect of parosmia is in progress.
iii. A case of excellent recovery from parosmia with significant improvement of olfactory function: After several years, one participant reported a “new normal” olfactory function, scoring 35 on the TDI test. This indicates regrowth of a broad range of healthy OSNs, yet two of the 52 aromas detected were still identified as triggers of parosmia: 2-methyl-3-furanthiol and one unidentified compound.
iv. A case of a functional ansomia with parosmia: At the other extreme one participant was within 3 months of onset and had a low TDI score (15) indicating functional anosmia. Only five aromas were detected, all scored as barely detectable, and only one, 2-furanmethanethiol, had parosmic character. Further investigations showed their parosmia was triggered more by different compounds present in onion and garlic.
v. A case of two distinct parosmic characters: one participant reported several distinct parosmic smells. One, which was described as plastic, chemical and burning rubber, was associated with sulfur compounds from Cluster A, and a second, described as sickly sweet, smoky and woody, was associated with pyrazines. Further investigation showed that onion and garlic gave a third parosmic character. These groupings are consistent with HCA clusters and a sub-theme emerging from the “AbScent Parosmia and Phantosmia Support” group on Facebook^6^.
vi. A case of sulfur triggers only: one participant attended just 4 weeks after onset of parosmia and was our most “fresh” parosmic. Their reactions at the GC-O were quite extreme when a trigger was encountered, with three thiols scoring 90-100 on the intensity scale. With a TDI score of 27, 20 GC-aromas were detected but only five of these were triggers, either thiols or disulfides. Pyrazines were detected but not reported as triggers. This is counter to any hypothesis which suggests that in the early days of parosmia, all (or many) compounds are triggers.
vii. A case of parosmia resulting from post-traumatic brain injury who reported the same molecular triggers as the post infectious participants. This participant found meat a far worse trigger than coffee, and this is reflected in detection of the meaty thiol rather than the coffee thiol.
viii. *3.5* Concentration *of stimulus* One participant assessed three different concentrations of coffee, spanning a factor of 10000. At the regular concentration, 11 aromas were detected, of which 5 were triggers. When the stimuli were diluted by 100, only 2 compounds were detected (barely detectable) but they still had the same character as in the regular coffee extract– one was still parosmic and the other was not. Dilution of the stimulus did not reverse the distortion, and this is backed up by many who still report parosmia character for weak and barely detectable aromas. When the stimuli were concentrated, those that were undistorted remained undistorted and there was no evidence that concentration of the stimuli could create more distortions. This is backed up by other participants reporting strong but undistorted aromas. On concentration, new aromas both distorted (4) and non-distorted (5) were detected as their concentrations became suprathreshold. Thus, for this parosmic, the concentration of the stimulus did not seem to change its character or determine its valence, which is often the case for perception of highly potent odorants, particularly sulfur compounds which on dilution can change from pungent to a pleasant fruity character.

### Summary

In summary, we have identified for the first time specific molecules which trigger parosmia. These experiments demonstrate that there is a common set of molecular triggers responsible for distortions and sense of disgust in coffee, and they also trigger parosmia in other chemically related foods. However, not all molecules in this set are triggers in all parosmic participants. These molecules tend to be potent, have very low detection thresholds and in isolation are neither distorted nor unpleasant for non-parosmic participants. However, odour activity is not the defining factor since (*E*)-β-damascenone has an exceptionally low odour threshold, is one of the more potent compounds in coffee^20^, and was always perceived as jammy and fruity by those parosmic participants who detected it. Most of the trigger molecules found in coffee belong to one of four distinct groups: thiols, pyrazines, disulfides, methoxypyrazines but there are no known odour receptors which are specific for the described trigger molecules. In addition, individual case studies suggest that parosmia symptoms are independent of olfactory function and parosmia may occur in patients with objectively normal olfactory function. Parosmic odour quality is not necessarily related to odour concentration. Recovery from parosmia can be associated with either improvement or stasis of olfactory function. As we stated in our hypothesis, in those with poor olfactory function, parosmia may be enhanced by a lack of contribution from other more desirable and less potent aroma compounds, but we found that the molecular triggers alone are the key drivers of parosmia and individually responsible for the perception of disgust. This explains why those with normal olfactory function, who also perceive the more desirable less potent aroma compounds, still experience parosmia.

## Discussion

Little is known about the pathophysiology of parosmia. Like the aetiologies of smell loss, both central and peripheral mechanisms have been proposed^21^ and can broadly be thought of as the central theory, the ephaptic theory and the mis-wiring theory. Although there is now some doubt about the role of the bulb in olfactory identification^22^, these theories take the standard model of glomerular activation pattern within the olfactory bulb as the motif of recognition within the CNS.

The central theory is based on the changes occurring in the integrative centres in the brain. A decrease in olfactory bulb volume ^23, 24^ and a significant loss of grey matter volume^25^ has been demonstrated in parosmic participants. Further evidence of central mechanisms has been published recently showing different fMRI activation patterns in parosmic participants compared to hyposmics^26^. Increased activation in the thalamus and the putamen was observed in the parosmic participants, the latter being of relevance since it is connected to the olfactory cortical networks and has been associated with the perception of disgust. Also, stronger activation was observed in the ventral striatum which is associated with odour valence. Whilst there is good evidence in humans for the central theory of parosmia, a purely central causation seems unlikely based on our evidence that parosmia is triggered by a group of highly specific molecules at the periphery.

The “mis-wiring” theory posits aberrant targeting of OSN to the glomerulus during regeneration from insult. This has been observed in mice with impaired olfactory function induced by ciliopathies^27-28^, physical lesioning^29-30^, and induced chemical degeneration^31^ but not yet in humans. However, it has been adopted as the likely mechanism for the perception of distorted olfactory percepts in parosmia. It is further suggested that the change in hedonic valence is due to broad activation of the olfactory bulb sending a disordered and unmoderated array of signals to the central neural processing system which invokes a strong sense of disgust. Our data neither support nor refute the mis-wiring hypothesis, but certainly place constraints on it. We have demonstrated the requirement to account for the non-stochastic nature of the OSNs involved in any proposed mis-wiring theory. Whilst mis-wiring is attributed to a loss of axonal pathfinding mechanisms^27^, in light of our results, this theory needs to explain why some OSNs are relaying the “correct” undistorted signal, whereas others are not, even early in the patient’s recovery.

The ephaptic theory summarised by Hawkes^32^ suggests that demyelination of the OSNs allow the activation of other, non-stimulated OSNs adjacent to the activated OSN by current flow in the extracellular fluid: “a form of short circuiting”. This too would result in a broader activation of the olfactory bulb and must be able to account for the non-random nature of the OSNs involved.

Of course, these theories are not mutually exclusive. Alterations in innervation of the glomeruli, or ephaptic activation of afferent nerves, and therefore the whole bulb, would result in the alteration of downstream central processing. The plasticity of this part of the pathway is much more limited. The “wiring” of the brain in recognising and acting on certain patterns of activation within the bulb remains after the peripheral insult. Therefore, a broader activation of the glomeruli would activate more odour object patterns within the cortex.

These hypotheses have to explain four characteristics: that parosmia arises almost uniquely in settings of widespread synchronous neuronal destruction either post infection or post traumatic brain injury, is triggered by one of a number of common odorants, is of novel odour character, and that this character is almost always unpleasant. Whereas the mis-wiring theory is consistent with the first, and the central theory may explain the novel odour character and the change in valence, it remains for us to determine why only a few potent molecules elicit such a strong parosmic response.

These trigger molecules share the trait of having an extremely low threshold in human olfaction, so they are detectable in very low concentrations. Such low olfactory thresholds may be attributed to a higher binding affinity for their respective olfactory receptors (ORs), but there are other possible explanations: higher rate of expression of specific ORs on the OSN cell surface, stronger activation of OSN depolarisation by the OR due to reduced habituation at the receptor or cellular levels, overrepresentation of that OSN within the mucosal OSN population, zonal expression (not demonstrated in humans) resulting in improved access to the nasal airstream, glomerular level or higher configurations resulting in greater salience of the glomerular activation, or the odorant could activate more than one olfactory receptor and the combined activation is summed as a lower detectable threshold within the bulb.

The common molecular structures, low odour thresholds and physiochemical grouping of the molecular triggers of parosmia strongly suggest that this is an olfactory receptor-level phenomenon, although we are unable to identify any specific olfactory receptors responsible from publicly available databases. The fact that some parosmic participants only report distortion with some of the groups suggests that there may be up to four separate olfactory receptors involved, or more when we consider other food and household items that elicit distortions. So why then these groups of OSNs? Several mechanisms could account for the role of specific receptors and their neurons.

1. These OSNs are regenerating because they are selectively damaged by the insult and others are preserved. This may account for the presence of parosmia in objectively normosmic volunteers, but it is unlikely that the insult from brain injury and viral infection would lead to the same preferential damage to particular OSNs.
2. The specific OR is predominant within the regenerating OSN population for one of two reasons. In one scenario, instead of a purely stochastic OR selection process in the olfactory mucosa, these “parosmic ORs” are preferentially selected for expression in OSNs either normally or just in the post-insult olfactory mucosa, thus increasing the number of these neurons in the olfactory mucosa as a whole. Alternatively, there is evidence in mice that activated OSNs have a longer lifespan^33^. Because of their ease of activation, these OSNs tend to survive longer and therefore make up a greater proportion of the overall OSN population.
3. The specific OR are not over-represented but merely more easily activated, so although many OSNs regenerate and aberrantly innervate the glomeruli, since only a few afferent neurons pass to each individual glomerulus, these few molecules are powerful enough to activate many glomeruli simultaneously at physiological odorant concentrations.
4. Since axon guidance is at least partially OR-dependant^34^ these specific OR-expressing OSNs could be more likely to demonstrate aberrant targeting of glomeruli in an as-yet unknown way.
5. It is possible that parosmia does not arise from the activation of the glomeruli per se, but the disruption of the network of interneurons, mitral, glial and tufted cells which is thought to act as a habituation and modulatory network in the bulb. Disruption of input innervation and sporadic re-innervation could cause feedback loops to interfere with the previous web of inhibition and promotion of signal at this level and this disordered activation is experienced as unpleasant.

Parosmia is a tetrapartite symptom: it is a triggered, short lived, altered smell sensation which almost universally elicits the basic emotion of disgust. Our finding that this is reliably triggered by a common group of low threshold odorants advances our understanding of this debilitating condition and constrains the pathophysiological hypothesis space.

That there are reliable molecular triggers of parosmia point to an olfactory receptor level pathology which agrees with the fact that the sensation is triggered by smells and follows the usual pattern of habituation and attenuation expected in an otherwise intact olfactory system. The selective regeneration of only a few OSNs also explains how the odour percept is altered (if the central dogma of glomerular activation patterns within the olfactory bulb is accepted). If only some of these glomeruli are activated out of the previous broader activation pattern recognised as coffee, this will be perceived as a novel smell.

What this does not explain is the presumably hypothalamic disgust response to this particular altered odour. The miswiring hypothesis posits that broader, unregulated patterns of glomerular activation are de novo perceived as unpleasant and disgusting, but this has not been demonstrated. Certainly, in the normal nose, novel smell percepts are not usually automatically disgusting, so the mere novelty of the percept is unlikely to be enough to explain this.

In this paper we identify the first common molecular triggers of human parosmia, characterised by their physiochemical properties and sharing a low odour threshold for humans. We demonstrate that parosmia is an olfactory dysfunction only partially correlated with olfactory loss, and provide evidence to support its arising in the periphery of the olfactory system. This information is vital to the understanding of the pathophysiology of this increasingly widespread condition and will be important in guiding further research and future therapies.

## Data Availability

All data freely available upon request

## Acknowledgements

The authors would like to acknowledge all those who participated in this study, Aidan Kirkwood for assistance with the participants, Peter Jackson for his assistance in sourcing and handling the faecal sample, and Professor Barry Smith for useful discussion and review of the manuscript.

## Authorship contribution

JP contributed to conception, acquisition, analysis and interpretation of data, manuscript draft and review; CK contributed to conception, participant management, data acquisition and review; SG contributed to conception, interpretation of data, manuscript draft and review (SG).

## Conflict of interest

The authors declare no conflict of interest.

## Methods

### Participants

This study (No 22/19) was approved by the University of Reading Research Ethics Committee. All parosmic participants were recruited via Facebook support groups or local ENT consultants, and non-parosmic participants from within the Department of Food and Nutritional Sciences or through private Facebook pages. The initial study was carried out with pre-COVID-19 parosmic participants (N=15) and non-parosmic participants (N=15) between October 2019 and March 2020. This was supplemented with post-COVID-19 parosmic participants (N=15) between July and September 2020. All volunteers completed a screening questionnaire (Supplementary Table S2) before attending a study day in the Olfaction Laboratory at the University of Reading. Selection was based on the participants listing coffee as a key trigger, and answering “often” at least once to two key questions which discriminate most efficiently between parosmic participants and those with quantitative olfactory disorders^35^:

1. Are odours that are pleasant to others, unpleasant to you? Never/rarely/often/always
2. Is the taste of food different to what you expect? Never/rarely/often/always

### Olfactory function

The bilateral olfactory function of all participants was assessed at the beginning of the day using the well-established and validated orthonasal psychophysical Sniffin’ Sticks test (Burghart, Wedel, Germany)^11^. Involving threshold (T), discrimination (D) and identification (I) tests, the resulting TDI score ranges from 0 to 48 with those scoring >30.3 classified as normosmic.

### Materials

Nescafé Original instant coffee sachets (Nestlé UK Ltd., York, UK) were used in this study. One box of instant Nescafé sachets (use by date August 2021) was purchased in September 2019 for use with the subjects between October 2019 and March 2020. A second box (use by date Oct 2022) was purchased in October 2020 to cross check the stability of the sachets over one year. Cocoa powder (Bournville, Cadbury, Bourneville, UK), skinless chicken breast fillet, smooth peanut butter (Tesco, Cheshunt, UK) and red bell peppers were purchased from a local supermarket. Faecal samples were kindly prepared under Class 2 conditions by members of the food and microbial science group at the University of Reading. The suppliers of all aroma standards are given in Table 2. Food safe high-grade mineral oil was purchased from Brandon Bespoke (Rochdale, UK), propylene glycol from Sigma (Poole, UK) and absorbent 3M PowerSorb P-110 paper from Merck (Gillingham, UK). A faecal sample was collected from a healthy donor who had not consumed antibiotics within the previous six months. For transportation to the laboratory the sample was held under anaerobic conditions, using an Oxoid Anaerogen sachet (Oxoid, Hampshire, UK), for up to two hours before being frozen at -20 °C.

### Extraction of coffee aroma

Fresh deionised water from a MilliQ system at 18.2 MΩ/cm resistivity was boiled in a kettle and 300 mL was added to the contents of the sachet (2.15 ± 0.05 g) in a 500 mL Duran bottle. The bottle was sealed, stirred for 2 min and an aliquot (3.0 ± 0.05 g) was transferred into vial. More concentrated extracts (contents of 1 sachet in 3 g boiling water) were also prepared for expert GC-O analysis and a detailed GC-MS analysis to aid identification of compounds. The vial was equilibrated at 55 °C for 20 min and a preconditioned triple phase solid phase microextraction (SPME) fibre (50/30 μm divinylbenzene/carboxen on polydimethylsiloxane (Supelco, Poole, UK)) was exposed to the headspace at 55 °C for 20 min prior to analysis by GC-O.

### Gas Chromatography-Olfactometry (GC-O)

After extraction, the SPME device was inserted into the injection port of an HP7890 GC from Agilent Technologies (Santa Clara, CA, USA) coupled to a Series II ODO 2 GC-O system (SGE, Ringwood, Victoria, Australia). The SPME fibre was desorbed in a split/splitless injection port held at 280 °C. The columns employed were either an Agilent HP-5 MSUi capillary (30 m, 0.25 mm i.d., 1.0 µm df) non-polar column or a Stabilwax^®^-DA (30 m, 0.25 mm i.d., 0.25 µm df) polar column (Restek, Bellefonte, PA, USA). The temperature gradients were set as follows: 40 °C for 2 min, then a rise of 5 °C/min up to 200 °C and 15 °C/min from 200 °C to 300 °C (or 250 °C for the polar column), and the final temperature held for a further 19 min. Helium was used as carrier gas (2 mL/min). At the end of the column, the flow was split 1:1 between a flame ionisation detector (kept at 250 °C) and a sniffing port using 2 untreated silica-fused capillaries of the same dimensions (1 m, 0.32 mm i.d.). The flow to the odour-port was diluted with a moist make up gas.

### Procedure at the odour-port

Subjects were familiarised with the instrument, instructed to breathe normally during the run, and advised that they could stop at any time, particularly if they felt dizzy or light-headed. As the aromas eluted from the column, 3 bits of information were requested from the subjects: an odour description, an odour intensity and an indication of whether the odour elicited a parosmic response. Since the description and identification of aromas in the absence of any other cues is difficult, all participants were presented with a flavour wheel (Supplementary Figure S1) before they started which they could use as a reference during the GC-O run. It had been developed by 2 experts who sniffed samples of the same coffee (both at regular strength and concentrated) by GC-O. The words were categorised into food and non-food, and colour coded for quick reference. This was of more use to non-parosmic participants, as parosmic participants found it hard to describe many of the aromas, even with the help of the flavour wheel. Many resorted to using the terms “new coffee”, “that parosmia smell”, “trigger number 1” or “trigger number 2”. As each aroma eluted, parosmic participants were prompted to highlight anything that had a parosmic character or trigger. Intensity was scored on a 158 mm horizontal general labelled magnitude scale (gLMS) with anchors at “barely detectable”, “weak”, “medium”, “strong”, “very strong” and “strongest imaginable” corresponding to intensity scores of 1.4, 6, 17, 35, 51 and 100 respectively (et al. 2004). This was chosen over the more common visual analogue scale to allow for instances where parosmic participants, in particular, wanted to extend upwards the range of scores. It is a logarithmic scale which better relates the psychophysics of perception to the concentration of the stimulus (Stevens’s Law). Time of elution was recorded manually by the researcher. All subjects carried out the GC-O of coffee twice, once before lunch and once after a 45 min lunch break. During the second run, the focus was on refining the descriptors as well as obtaining a duplicate intensity rating. During the first run, the subjects recorded the descriptors in their own words, prompted only by the flavour wheel, whereas during the second run, there was more discussion between the researcher and the subject, to verify the odour character and identity of the compound eluting.

### Gas chromatography-mass spectrometry (GC-MS)

An extract from a coffee prepared with one sachet in 3 mL of boiling water was extracted as above and analysed by GC-MS to aid identification of aroma compounds detected by GC-O and confirm their presence in the coffee extract. A7890A Gas Chromatograph coupled to a 5975C series GC/MSD from Agilent was used, equipped with either of the columns described above. The oven was held at 40 °C for 2 min, increased from 40 °C to 250 °C at a rate of 4 °C/min and then kept constant at 250 °C for 5 min. Helium was the carrier gas at a flow rate of 1.2 mL/min. Mass spectra were recorded in electron impact mode at an ionisation voltage of 70 eV and source temperature of 220 °C. A scan range of m/z 20-300 with a scan time of 0.69 s was employed and the data were controlled and stored by the ChemStation software (Agilent, Santa Clara, CA).

### Identification of odour-active compounds

Linear retention indices were calculated by comparison with the retention times of C_6_-C_25_ n-alkane series analysed on the same day using the same conditions as for sample analyses (Supplementary Table S3). Aromas eluting from the GC-O were identified by comparing their LRIs, mass spectra and the odour as described by the experts with those of authentic compounds on two columns of different polarity. Mass spectral libraries, such as NIST 2011 and Inramass (INRA, France), were used for primary identification of compounds in the coffee extract using ChemStation software (Agilent, Santa Clara, CA). In most cases, authentic compounds were analysed using the same chromatographic method to confirm their identity by comparison of their mass spectra, LRI, and odour quality. Identification was confirmed by GC-MS on a Stabilwax column. For compounds at concentrations below the detection limit of the GC-MS, odour character and LRI were used.

### Confirmation of molecules as trigger or non-trigger

Three parosmic participants returned to assess coffee on a polar column to confirm the identity of trigger compounds. Once identified, selected trigger compounds were also presented to 2 parosmic participants in dilute form to verify their parosmic character, using the sample preparation protocol described for the European test of olfactory capabilities^36^. Aroma chemicals were diluted in mineral oil or propylene glycol and applied to small discs (5 mm diameter) of absorbent paper in vials which were presented to the participants. They were asked to sniff the vial and indicate whether each compound released “that parosmia smell” which they had described previously.

### Additional samples

All additional samples were prepared as for coffee with the following modifications. Cocoa: 3g of cocoa powder was dissolved in 10 g boiling water, stirred and a 3 g aliquot was used for extraction. Meat: a lean breast fillet, thickness 1 cm was grilled for 3 min on either side using a Cuisinart grill (Stamford, CT) set on high. Finely chopped meat (3 g) was used for extraction. A 50:50 slurry of peanut butter (3 g) was used for extraction. Finely diced red pepper (3 g) was extracted at 40 °C prior to desorption. The faecal sample was thawed, mixed with an equal weight of water, and 3 g transferred to an SPME vial Chromatography conditions for all samples remained the same as for coffee.

### Statistics

Analysis of variance was carried out on age and TDI scores to determine whether there were significant differences between pre-COVID-19 parosmic participants, post-COVID-19 parosmic participants and non-parosmic participants. Post hoc pairwise comparisons were performed using Fishers LSD at p=0.05. Agglomerative hierarchical clustering (AHC) using Ward’s Euclidean distance was carried out on intensity data for the 17 compounds most frequently identified in coffee as molecular triggers of parosmia. All statistical analyses were carried out using XLSTAT statistical and data analysis solution (Addinsoft 2020).

---

**Table S1.**
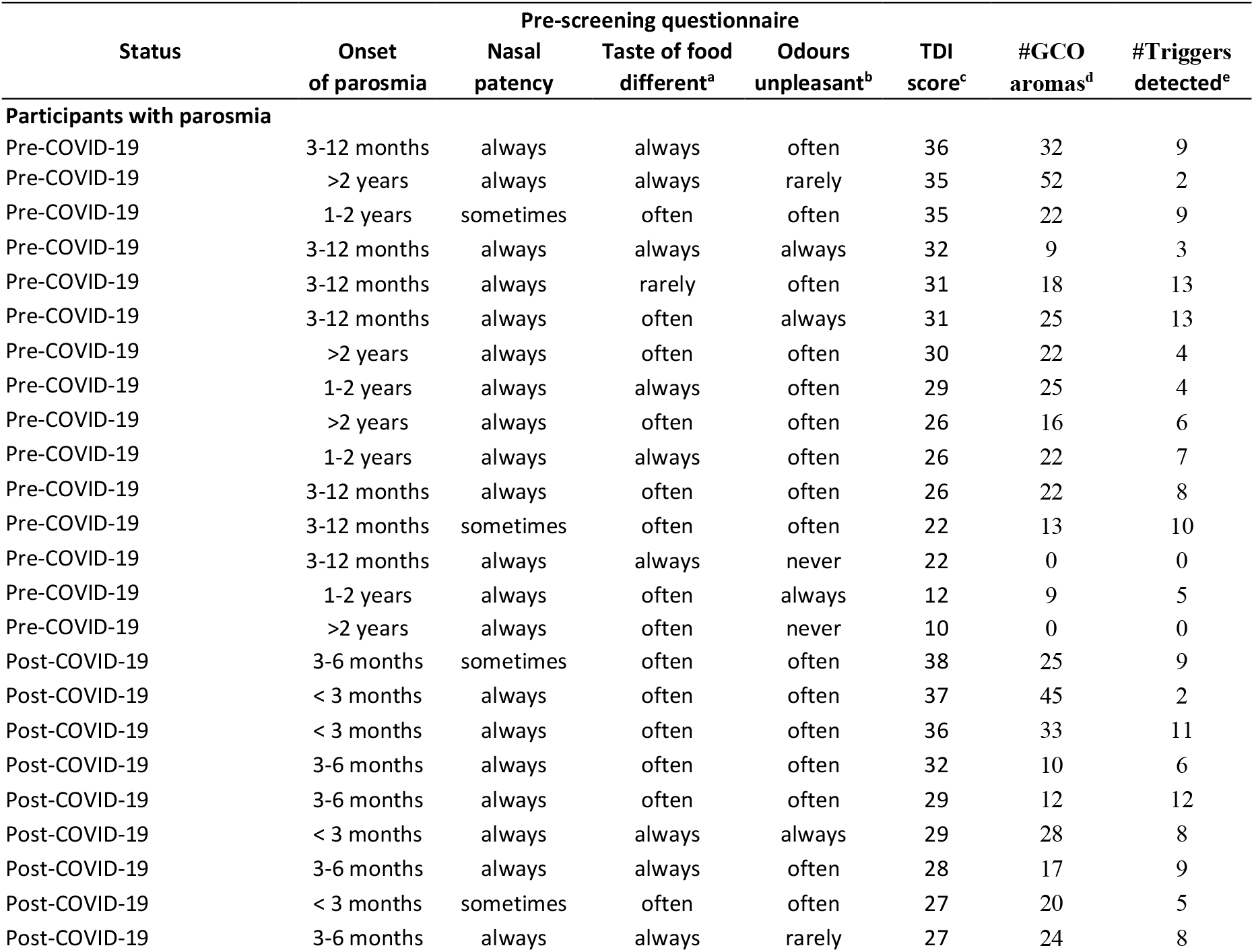
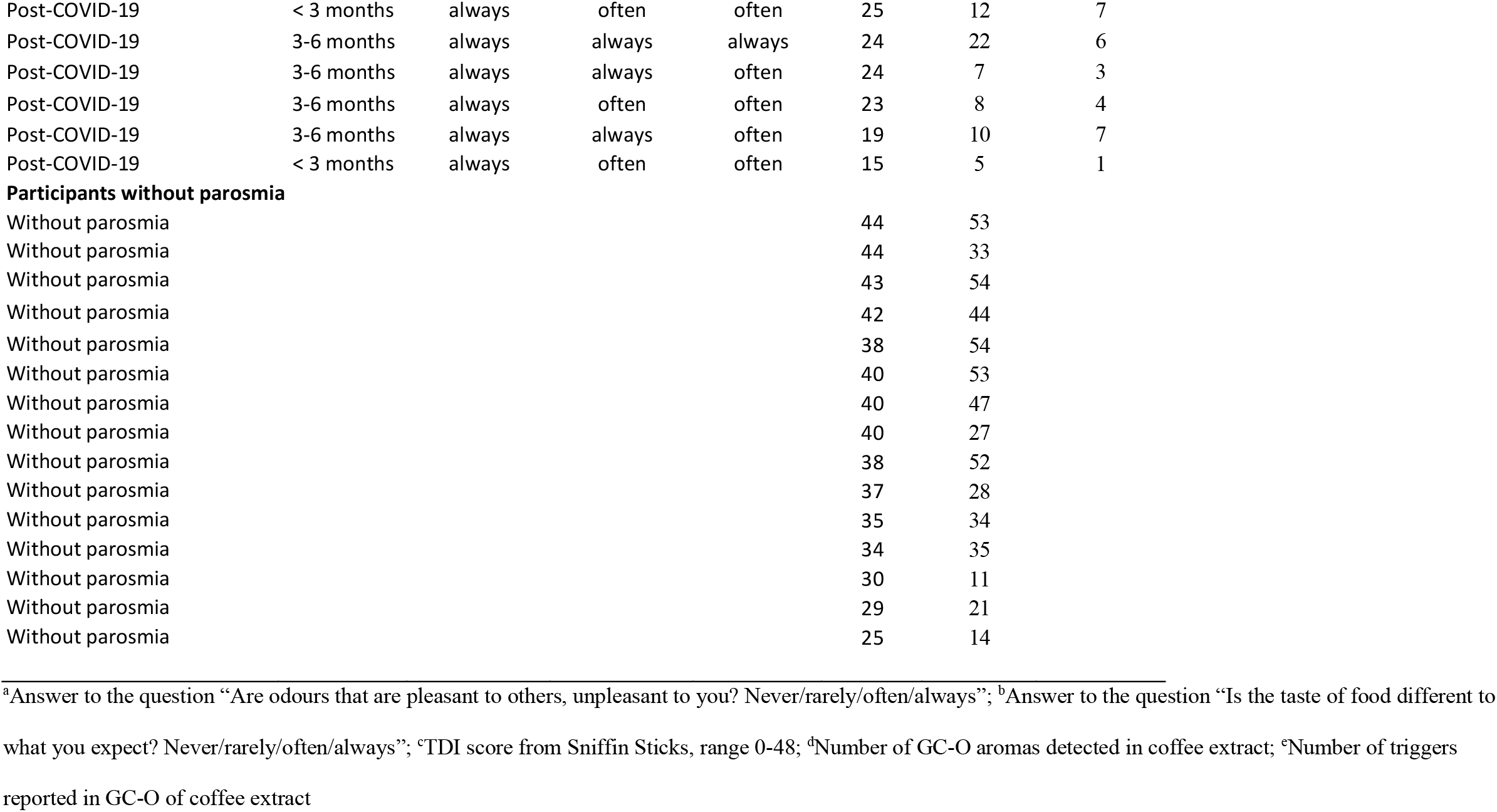
Participant Data: Demographics, answers to pre-screening questionnaire, TDI Scores and GC-O scores

**Table S2.**
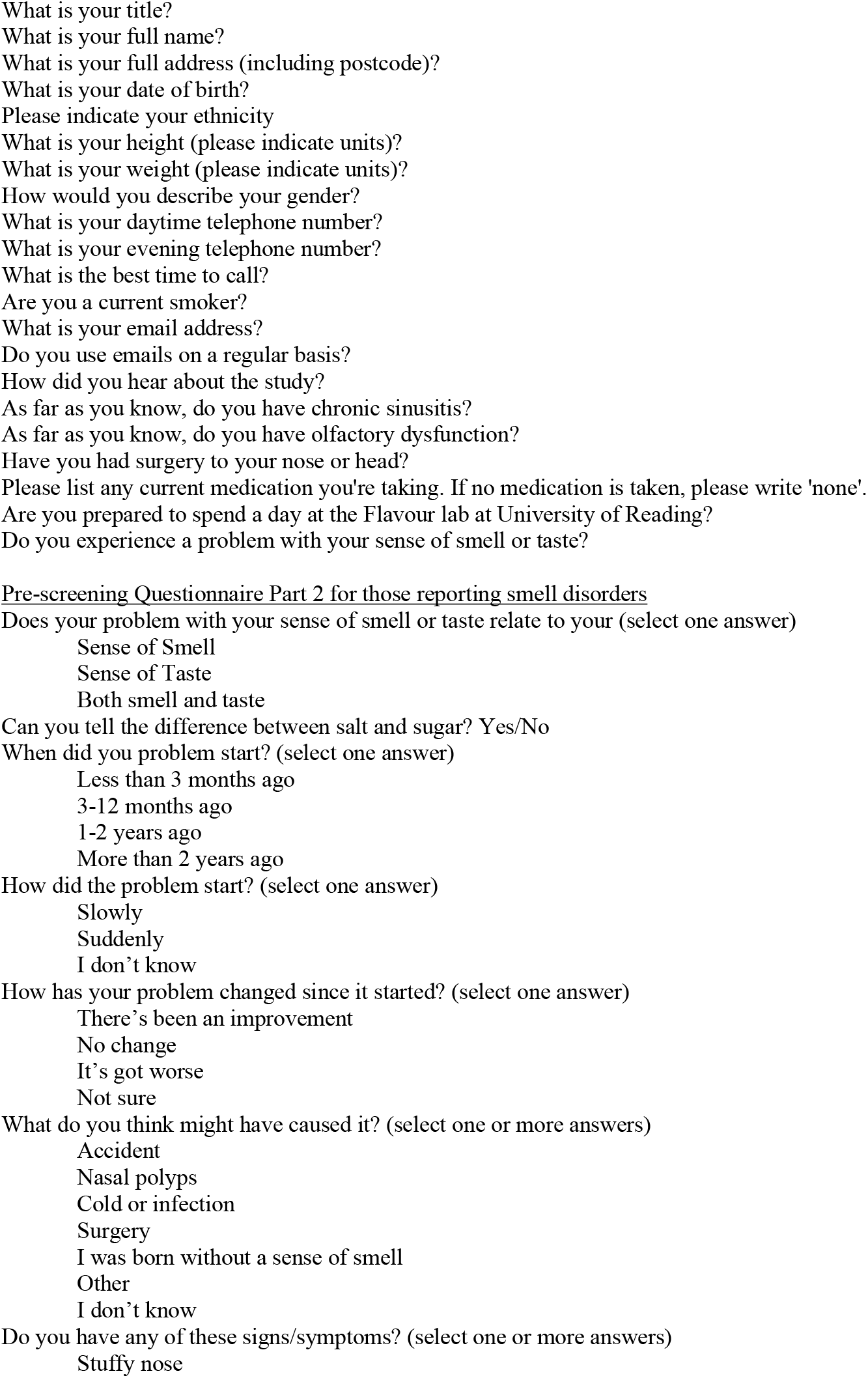
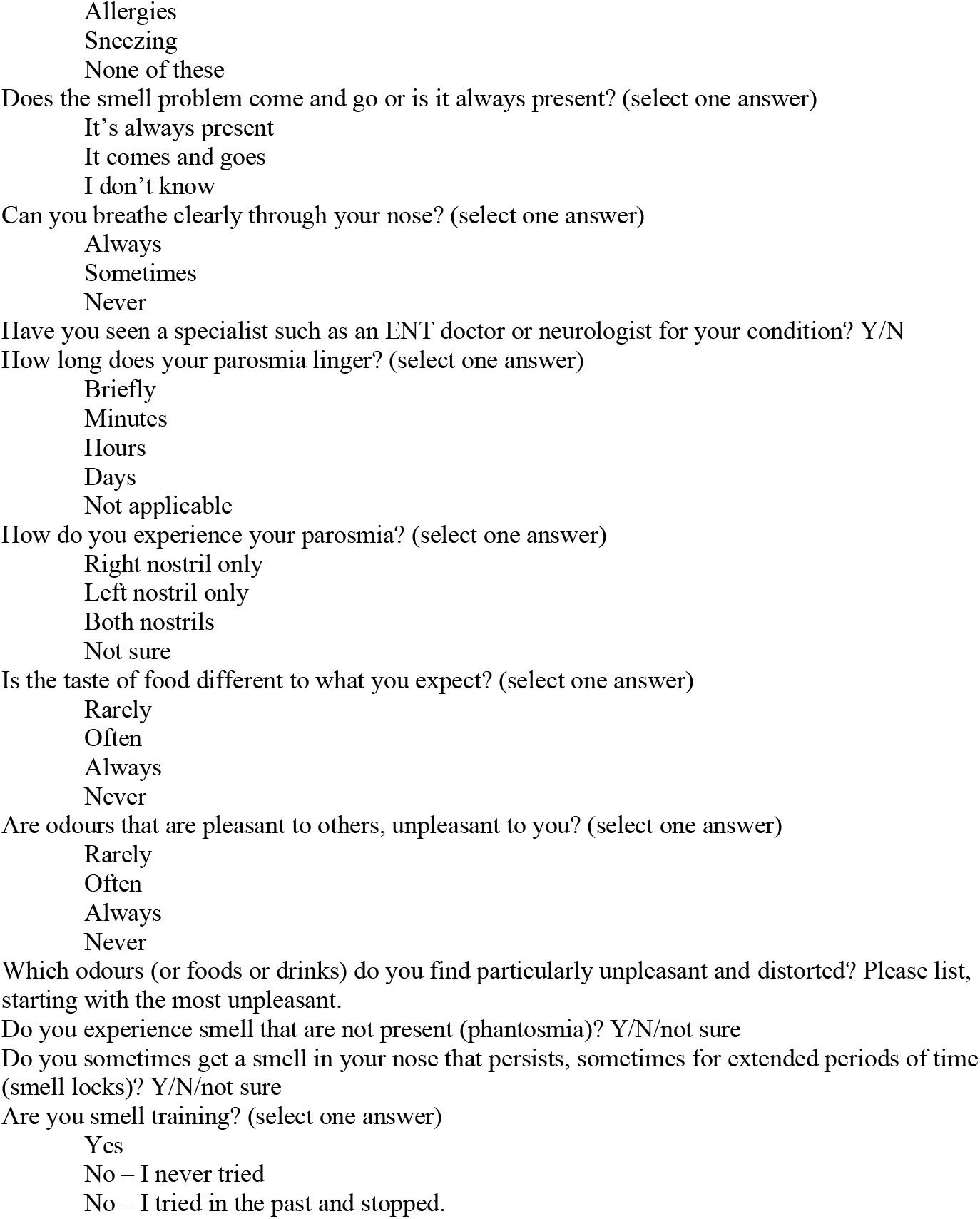
Pre-screening Questionnaire Part 1

**Table S3.**
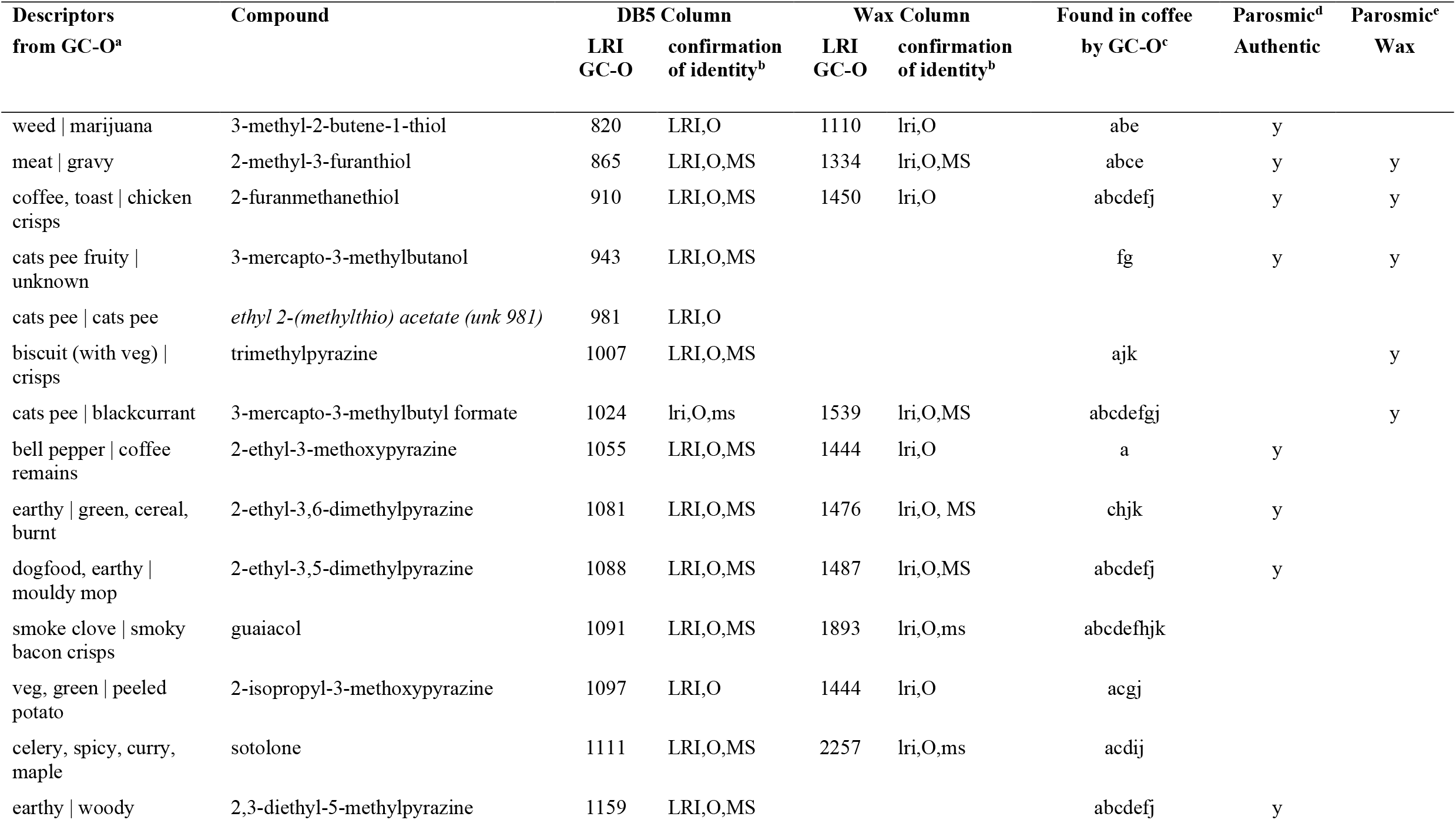
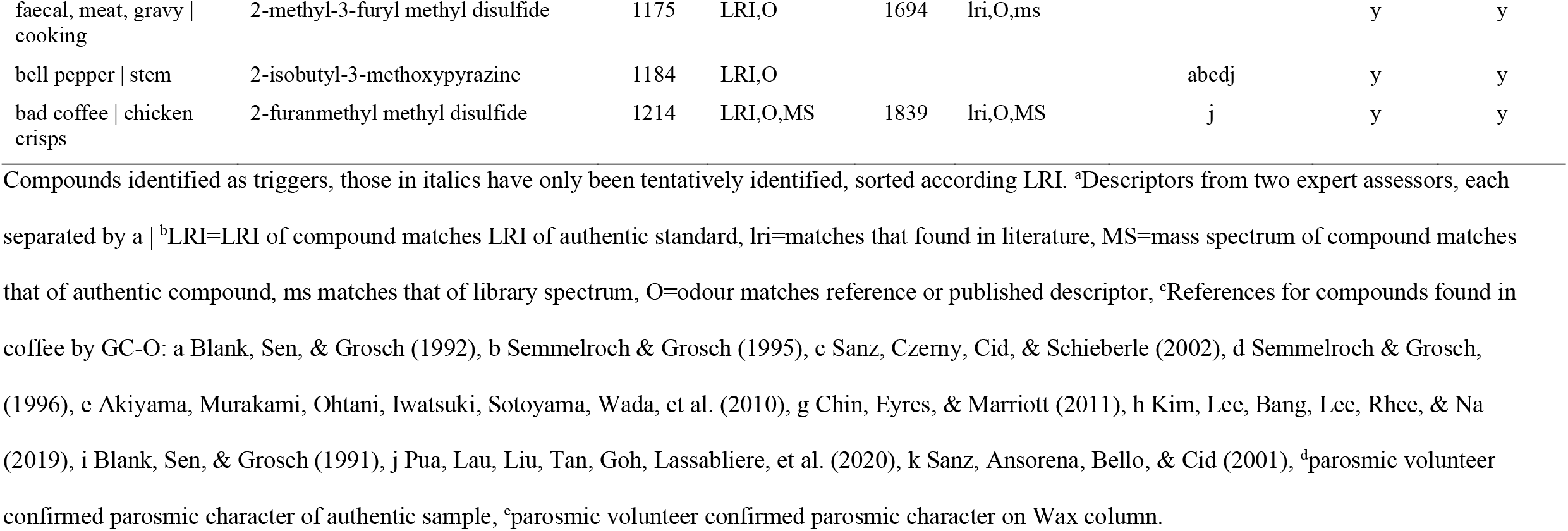
Confirmation of Identity of Molecular Triggers of Parosmia

**Figure S1.**
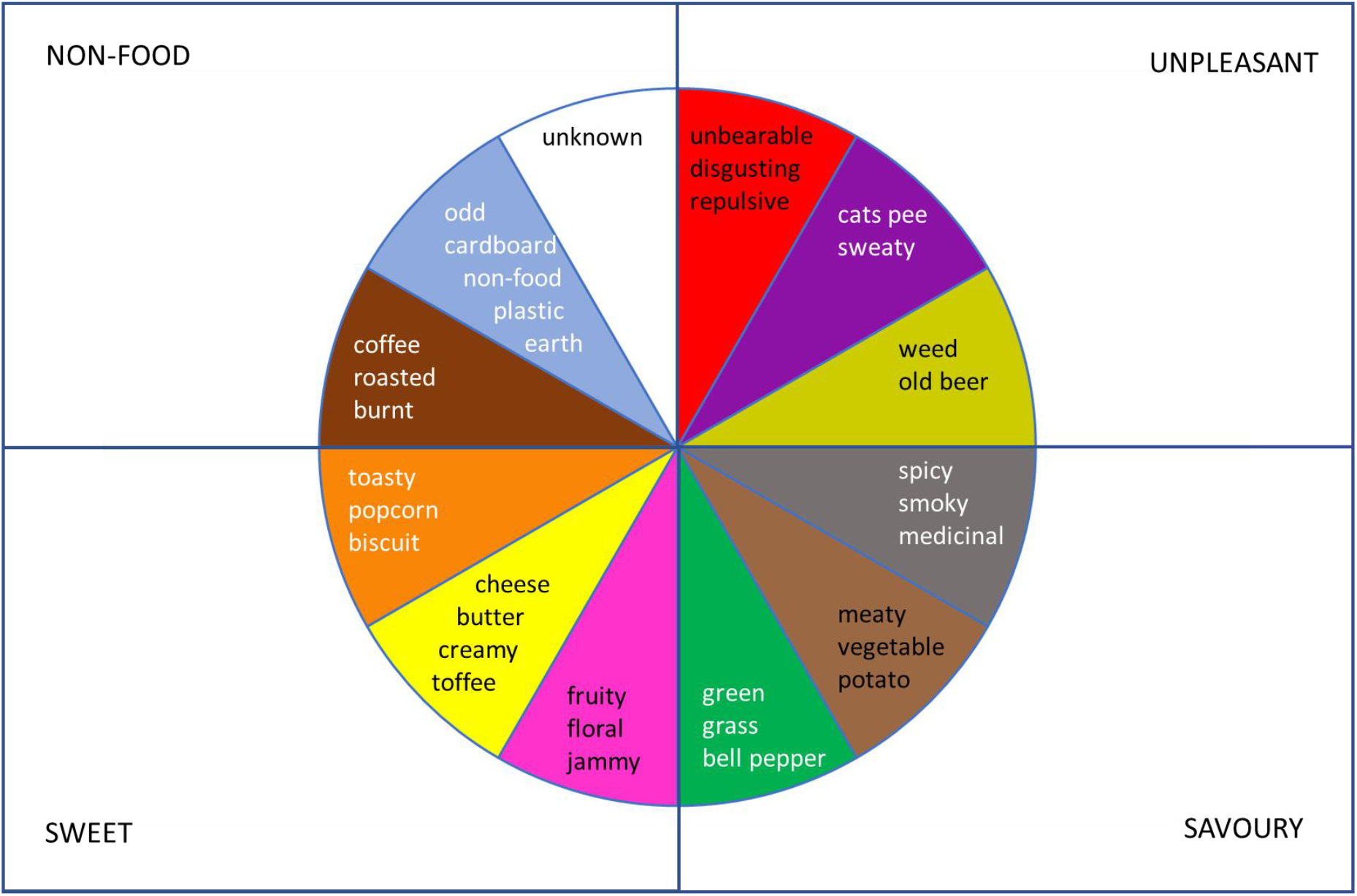
Flavour wheel provided to all participants during GC-O session

## Notes

### Competing Interest Statement

The authors have declared no competing interest.

### Funding Statement

No external funding was received.

### Author Declarations

Unniversity of Reading Research Ethics Committee

## References

1. Boscolo-Rizzo, P.; Polesel, J.; Spinato, G.; Menegaldo, A.; Fabbris, C.; Calvanese, L.; Borsetto, D.; Hopkins, C.; Predominance of an altered sense of smell or taste among long-lasting symptoms in patients with mildly symptomatic COVID-19. Rhinology 2020, 58 (5), 524–525.

2. Gane, S. B.; Kelly, C.; Hopkins, C.; Isolated sudden onset anosmia in COVID-19 infection. A novel syndrome? Rhinology 2020, 58 (3), 299–301.

3. Boscolo-Rizzo, P.; Menegaldo, A.; Fabbris, C.; Spinato, G.; Borsetto, D.; Vaira, L. A.; Calvanese, L.; Pettorelli, A.; Sonego, M.; Frezza, D.; Bertolin, A.; Cestaro, W.; Rigoli, R.; Tirelli, G.; Mosto, M. C. D.; Menini, A.; Polesel, J.; Hopkins, C.; High prevalence of long-term psychophysical olfactory dysfunction in patients with COVID-19. medRxiv 2021, 2021.01.07.21249406.

4. WHO WHO Coronavirus Disease (COVID-19) Dashboard. (accessed 12th December 2020).

5. Bonfils, P.; Avan, P.; Faulcon, P.; Malinvaud, D.; Distorted odorant perception - Analysis of a series of 56 patients with parosmia. Archives of Otolaryngology-Head & Neck Surgery 2005, 131 (2), 107–112.

6. Parker, J. K.; Kelly, C. E.; Smith, B.; Hopkins, C.; S., G., An analysis of patients’ perspectives on qualitative olfactory dysfunction using social media. medRxiv 2020.12.30.20249029; doi: https://doi.org/10.1101/2020.12.30.20249029 2021.

7. Burges Watson, D. L., Campbell, M.; Hopkins, C.; Smith, B.; Kelly, C.; Deary, V.; Altered Smell and Taste: anosmia, parosmia and the impact of long Covid-19. medRxiv 2020, 2020.11.26.20239152.

8. Croy, I.; Nordin, S.; Hummel, T.; Olfactory Disorders and Quality of Life-An Updated Review. Chemical Senses 2014, 39 (3), 185–194.

9. Kohli, P.; Soler, Z. M.; Nguyen, S. A.; Muus, J. S.; Schlosser, R. J.; The Association Between Olfaction and Depression: A Systematic Review. Chemical Senses 2016, 41 (6), 479–486.

10. Keller, A.; Malaspina, D.; Hidden consequences of olfactory dysfunction: a patient report series. BMC ear, nose, and throat disorders 2013, 13 (1), 8–8.

11. Hummel, T.; Sekinger, B.; Wolf, S. R.; Pauli, E.; Kobal, G.; ‘Sniffin’ Sticks’: Olfactory performance assessed by the combined testing of odor identification, odor discrimination and olfactory threshold. Chemical Senses 1997, 22 (1), 39–52.

12. Hummel, T.; Kobal, G.; Gudziol, H.; Mackay-Sim, A.; Normative data for the “Sniffin’ Sticks” including tests of odor identification, odor discrimination, and olfactory thresholds: an upgrade based on a group of more than 3,000 subjects. European Archives of Oto-Rhino- Laryngology 2007, 264 (3), 237–243.

13. Czerny, M.; Christlbauer, M.; Christlbauer, M.; Fischer, A.; Granvogl, M.; Hammer, M.; Hartl, C.; Hernandez, N. M.; Schieberle, P.; Re-investigation on odour thresholds of key food aroma compounds and development of an aroma language based on odour qualities of defined aqueous odorant solutions. Eur. Food Res. Technol. 2008, 228 (2), 265–273.

14. Rychlik, M.; Schieberle, P.; Grosch, W.; Compilation of ordor thresholds, odor qualities and retention indices of key food odorants. Deutsche Forschungsanstalt fur Lebensmittelchemie: Garching, Germany: 1998.

15. Blank, I.; Sen, A.; Grosch, W.; Potent odorants of the roasted powder and brew of Arabica coffee. Z. Lebensm.-Unters. Forsch. 1992, 195 (3), 239–45.

16. Fors, S.; Sensory properties of volatile Maillard reaction products and related compounds: a literature review. ACS Symp. Ser. 1983, 215, 185–286.

17. Liu, X.; Su, X.; Wang, F.; Huang, Z.; Wang, Q.; Li, Z.; Zhang, R.; Wu, L.; Pan, Y.; Chen, Y.; Zhuang, H.; Chen, G.; Shi, T.; Zhang, J.; ODORactor: a web server for deciphering olfactory coding. Bioinformatics 2011, 27 (16), 2302–2303.

18. Sanz, G.; Thomas-Danguin, T.; Hamdani, E. H.; Le Poupon, C.; Briand, L.; Pernollet, J.-C.; Guichard, E.; Tromelin, A.; Relationships between molecular structure and perceived odor quality of ligands for a human olfactory receptor. Chemical Senses 2008, 33 (7), 639–653.

19. Moore, J. G.; Straight, R. C.; Osborne, D. N.; Wayne, A. W.; Olfactory, gas- chromatographic and mass-spectral analyses of fecal volatiles traced to ingested licorice and apple. Biochemical and Biophysical Research Communications 1985, 131 (1), 339–346.

20. Sanz, C.; Czerny, M.; Cid, C.; Schieberle, P.; Comparison of potent odorants in a filtered coffee brew and in an instant coffee beverage by aroma extract dilution analysis (AEDA). Eur. Food Res. Technol. 2002, 214 (4), 299–302.

21. Leopold, D.; Distortion of olfactory perception: Diagnosis and treatment. Chemical Senses 2002, 27 (7), 611–615.

22. Weiss, T.; Soroka, T.; Gorodisky, L.; Shushan, S.; Snitz, K.; Weissgross, R.; Furman-Haran, E., Dhollander, T.; Sobel, N.; Human Olfaction without Apparent Olfactory Bulbs. Neuron 2020, 105 (1), 35-+.

23. Mueller, A.; Rodewald, A.; Reden, J.; Gerber, J.; von Kummer, R.; Hummel, T.; Reduced olfactory bulb volume in post-traumatic and post-infectious olfactory dysfunction. Neuroreport 2005, 16 (5), 475–478.

24. Rombaux, P.; Mouraux, A.; Bertrand, B.; Nicolas, G.; Duprez, T.; Hummel, T.; Olfactory function and olfactory bulb volume in patients with postinfectious olfactory loss. Laryngoscope 2006, 116 (3), 436–439.

25. Bitter, T.; Siegert, F.; Gudziol, H.; Burmeister, H. P.; Mentzel, H. J.; Hummel, T.; Gaser, C.; Guntinas-Lichius, O.; Gray matter alterations in parosmia. Neuroscience 2011, 177, 177–182.

26. Iannilli, E.; Leopold, D. A.; Hornung, D. E.; Hummel, T.; Advances in Understanding Parosmia: An fMRI Study. Orl-Journal for Oto-Rhino-Laryngology Head and Neck Surgery 2019, 81 (4), 185–192.

27. Green, W. W.; Uytingco, C. R.; Ukhanov, K.; Kolb, Z.; Moretta, J.; McIntyre, J. C.; Martens, J. R.; Peripheral Gene Therapeutic Rescue of an Olfactory Ciliopathy Restores Sensory Input, Axonal Pathfinding, and Odor-Guided Behavior. Journal of Neuroscience 2018, 38 (34), 7462–7475.

28. Tadenev, A. L. D.; Kulaga, H. M.; May-Simera, H. L.; Kelley, M. W.; Katsanis, N.; Reed, R. R.; Loss of Bardet-Biedl syndrome protein-8 (BBS8) perturbs olfactory function, protein localization, and axon targeting. Proceedings of the National Academy of Sciences of the United States of America 2011, 108 (25), 10320–10325.

29. Christensen, M. D.; Holbrook, E. H.; Costanzo, R. M.; Schwob, J. E.; Rhinotopy is disrupted during the re-innervation of the olfactory bulb that follows transection of the olfactory nerve. Chemical Senses 2001, 26 (4), 359–369.

30. Costanzo, R. M.; Rewiring the olfactory bulb: Changes in odor maps following recovery from nerve transection. Chemical Senses 2000, 25 (2), 199–205.

31. St John, J. A.; Key, B.; Axon mis-targeting in the olfactory bulb during regeneration of olfactory neuroepithelium. Chemical Senses 2003, 28 (9), 773–779.

32. Hawkes, C.; Parosmia: treatment, mechanism, and types. British Medical Journal 2020, 371.

33. Watt, W. C.; Sakano, H.; Lee, Z. Y.; Reusch, J. E.; Trinh, K.; Storm, D. R.; Odorant stimulation enhances survival of olfactory sensory neurons via MAPK and CREB. Neuron 2004, 41 (6), 955–967.

34. Schwarting, G. A.; Henion, T. R.; Olfactory axon guidance: the modified rules. Journal of neuroscience research 2008, 86 (1), 11–7.

35. Landis, B. N.; Frasnelli, J.; Croy, I.; Hummel, T.; Evaluating the Clinical Usefulness of Structured Questions in Parosmia Assessment. Laryngoscope 2010, 120 (8), 1707–1713.

36. Thomas-Danguin, T.; Rouby, C.; Sicard, G.; Vigouroux, M.; Farget, V.; Johanson, A.; Bengtzon, A.; Hall, G.; Ormel, W.; De Graaf, C.; Rousseau, F.; Dumont, J. P.; Development of the ETOC: A European test of olfactory capabilities. Rhinology 2003, 41 (3), 142–151.

37. Karahadian, C.; Johnson, K. A.; Analysis of headspace volatiles and sensory characteristics of fresh corn tortillas made from fresh masa dough and spray-dried masa flour. J. Agric. Food Chem. 1993, 41 (5), 791–9.

